# An assessment of a novel franchise model for addressing the challenge of sustainable water access in resource-poor settings

**DOI:** 10.1101/2025.10.26.25338842

**Authors:** Martin Muchangi, Daniel Kurao, Kennedy Omwaka, Kioko Kithuki, Tara Klijn, Maarten Kuijpers, Gilbert Wangalwa, Kimani Karuga, Sammy Kemboi, Alvin Miheso

## Abstract

Access to safe drinking water remains a critical challenge in many resource-poor settings, especially in rural and peri-urban areas. In Kenya, despite constitutional guarantees for water access, millions still rely on unsafe water sources, contributing to high disease burdens. To address this gap, Amref Health Africa, in partnership with MegaGroup Netherlands, piloted the WaterStarters franchise model. This innovative model sought to improve sustainable water access by empowering local private and community-based organizations to manage water systems through a hybrid financing approach. The study aimed to evaluate the effectiveness, financial viability, and scalability of the WaterStarters model in enhancing water access in Kajiado County. Using a mixed-methods design, the study drew on survey data from n=1,209 households, key informant interviews, and focus group discussions across five pilot sites. The findings indicated that the model improved access to safe, affordable, and reliable water, with notable benefits in hygiene, reduced disease incidence, and time savings for households, particularly benefiting women and girls. However, financial sustainability was mixed, with only one pilot site achieving positive financial returns under the initial model. The study concluded that the WaterStarters franchise model holds promise for addressing water access gaps in underserved regions, especially when combined with appropriate financial restructuring. It recommended scaling the model with adjustments to financing, emphasizing local ownership, and integrating technology for improved service delivery. Continued investment in capacity building and governance structures is critical for long-term success and sustainability.

## Introduction

According to the WHO/UNICEF Joint Monitoring Programme for Water Supply, Sanitation and Hygiene, approximately 2.2 billion people globally lacked access to safely managed drinking water services as of 2022. This figure includes 1.5 billion individuals relying on basic services, 292 million with limited services, 296 million using unimproved sources, and 115 million who still depend on surface water for drinking. Alarmingly, 80% of those without basic water services live in rural areas. From 2015 to 2020, the global use of safely managed drinking water services increased by an average of 0.63 percentage points annually. However, to achieve universal coverage by 2030, the current rate of progress must accelerate fourfold [1].

Globally, diarrhoeal disease remains a major public health challenge, particularly for young children. It is the third leading cause of death among children aged 1–59 months, despite being both preventable and treatable [2]. Each year, diarrhoea claims the lives of approximately 444,000 children under the age of five, translating to over 1,200 child deaths per day [1]. Beyond mortality, diarrhoea is a key contributor to malnutrition in children. A significant proportion of the diarrhoeal disease burden is attributed to inadequate access to safe drinking water, sanitation, and hygiene (WASH), which also drives the spread of water- and hygiene-related neglected tropical diseases (NTDs) such as trachoma, schistosomiasis, and Guinea worm disease [3][4]. Inadequate WASH services disproportionately impact women and girls, exposing them to gender-based violence, diminished dignity, and lost productivity [4]. In many remote settings, they endure unsafe and time-consuming journeys - often exceeding 30 minutes per trip - to access clean and safe water [1][5].

In Kenya, access to safe water and sanitation remains a critical public health and development challenge. Approximately 39.4% of the population - about 18.7 million people - lack access to safe drinking water [1]. The situation is especially dire in rural areas, where nearly 24.1% of residents rely on unsafe sources such as rivers, lakes, and ponds [10]. National estimates indicate that only about 59% of Kenyans have access to safe water, reflecting persistent inequalities and underscoring the urgent need for investment in WASH infrastructure and governance. The United Nations classifies Kenya as a chronically water-scarce country, with natural water availability standing at just 647 cubic meters per capita per year - far below the global benchmark of 1,000 cubic meters [11]. This water scarcity contributes to widespread vulnerability, especially in rural and informal urban settlements where basic water and sanitation services are either lacking or poorly maintained.

Similarly, sanitation coverage has deteriorated, declining by five percentage points from 34% in 2000 to 29% in 2021, indicating a worsening public health challenge [1]. Approximately 50% of rural households have no toilet facilities at all, and where they exist, they are often unhygienic. In urban areas, up to 50% of the population lives in slum environments with deplorable sanitation conditions. Schools are also affected, with an average of one latrine for every 100 pupils, compared to the recommended maximum of 40 pupils per latrine [6].

These deficits in WASH infrastructure have significant health consequences. In Kenya, around 80% of hospital visits are due to preventable diseases, and about half of these are water, sanitation, and hygiene-related [7]. Additionally, over 90% of water and sanitation-related disease outbreaks occur in rural areas, and more than three-quarters of Kenya remains vulnerable to disasters such as floods, droughts, and cholera outbreaks. Addressing Kenya’s WASH challenges is therefore essential not only for improving health outcomes but also for achieving broader goals of equity, resilience, and sustainable development. To its credit, Kenya has developed a robust policy and legislative framework for WASH service delivery, anchored in Article 43 of the Constitution, which guarantees the right to clean water and sanitation. Legal instruments such as the Water Act (2016), National Water Master Plan 2030, and Environmental Sanitation and Hygiene Policy (2016–2030) reflect alignment with Sustainable Development Goal 6 (SDG 6), while providing guidance for integrated resource management, regulation, and service expansion. However, implementation remains the critical bottleneck. Persistent underfunding, fragmented institutional coordination, and limited technical capacity continue to undermine efforts to deliver WASH services at scale and fully realize the right to safe water and sanitation [8][5].

### The WaterStarters model

The WaterStarters model represents a transformative shift in rural water service delivery, moving away from traditional donor-dependent approaches toward a locally driven, franchise-based model that emphasizes community ownership, financial co-investment, and sustainability. At its core, the model empowers local franchisees (typically private operators or community-based organizations) to co-invest in and assume full responsibility for the management and operations of water systems. Its innovative hybrid financing mechanism requires a 15% upfront contribution from franchisees, with the remaining 85% financed through a blend of grant and recoverable investment (42.5% each), repaid over four years at 10% interest on a reducing balance. Technological integration is another defining feature, with prepaid water meters, solar-powered pumping systems, and a real-time digital monitoring dashboard deployed to enhance efficiency, transparency, and service reliability. The model’s long-term vision is to recover enough capital through repayments to build a revolving fund that will fuel its scale-up, with a goal of reaching 1.5 million people by 2030. As of April 2025, the model had been piloted in seven sites across Kajiado County.

### Study aims and objectives

This study aimed to assess the effectiveness, financial viability, and scalability of the WaterStarters franchise model as an innovative, community-based solution for expanding sustainable water access in resource-constrained settings. To achieve this aim, the study pursued the following objectives:

1. Evaluate the operational effectiveness of the WaterStarters model in enhancing access to safe, reliable, and affordable water, and its impact on health outcomes, household well-being, gender equity, and climate resilience.
2. Assess the financial performance and sustainability of the franchise model, including cost-recovery mechanisms, revenue generation, and the feasibility of the hybrid financing structure across diverse settings.
3. Examine the governance, technical, and contextual factors shaping implementation success, and identify the conditions, enablers, and barriers that affect the scalability of the WaterStarters model.

### Research Questions

The study was guided by the following overarching research questions:

1. How effective is the WaterStarters model in delivering continuous, safe, and equitable water services, and what are the resulting health, social, and economic outcomes - particularly for women, girls and vulnerable groups?
2. To what extent is the franchise-based model financially viable and sustainable, and how do financing parameters (e.g., loan-to-grant ratio, tariff structures, franchisee contributions) influence business performance and long-term profitability?
3. What technical, operational, and contextual factors explain variation in performance across sites, and how can risk management and governance structures be optimized to improve implementation outcomes?
4. What conditions are necessary to scale the WaterStarters model, and what adaptations are required to ensure its success in different geographic and market contexts?

## Materials and methods

### Study setting

The study was conducted in Kajiado County, Kenya, an arid and semi-arid region in the country’s southwest, bordering Nairobi and Tanzania. With a population projected to reach 1.3 million by 2025, the county faces chronic water scarcity, worsened by erratic rainfall and prolonged droughts. Only 66% of residents use improved water sources, while sanitation coverage stands at 56%, with 32% of the population practicing open defecation. Urban areas suffer from unreliable piped water and unregulated liquid and solid waste management, while rural communities often rely on unsafe sources located over 5 km away [9].

### Study population

The study population comprised both direct beneficiaries and key implementation stakeholders of the WaterStarters Programme across five project sites in Kajiado County: Kumpa, Orinie, Naserian, Kima, and Kibiko. Geographical coverage in each site was defined through participatory mapping with local focal persons to ensure inclusion of respondents directly served by or affected by the water projects. This approach enhanced the contextual relevance, accuracy, and utility of the findings.

The quantitative component targeted adult household members residing within the catchment areas, including those using water from the project as well as those relying on alternative sources. Households were further disaggregated by demographic and socio-economic characteristics such as age, gender, and primary livelihood activity to capture a wide range of user experiences and equity-related outcomes. The qualitative component engaged a diverse set of stakeholders involved in or impacted by the programme, including franchisees, public health officers, county officials, community health promoters (CHPs), water management committees, school staff, community leaders, and project implementing partners.

### Research Design

The study adopted a descriptive cross-sectional design grounded in a mixed-methods approach. This enabled the collection of both quantitative and qualitative data to capture the operational performance, financial sustainability, and social impact of the WaterStarters Programme across five implementation sites. The design allowed for in-depth analysis of programme outcomes at a single point in time, while triangulation of multiple data sources enhanced the credibility of findings.

### Sample size and sampling technique

The study used a two-pronged sampling approach. For the quantitative component, a cluster random sampling method was used. Households were selected within identified villages in each site using every third household interval, guided by a pen-toss random walk method. The culturally specific unit of an *olmarei* (wife’s dwelling in a polygamous homestead) was treated as a household. The final sample included 1,217 households: Kumpa (247), Orinie (247), Kima (253), Naserian (178), and Kibiko (292). For the qualitative component, purposive sampling was used to select participants for 14 key informant interviews and 20 focus group discussions. Respondents were chosen based on their relevance to programme implementation and knowledge of local water service dynamics.

### Data collection instruments

Quantitative data were collected using a structured household questionnaire programmed on KoBoToolbox and administered via mobile phones. The survey captured information on water access, usage, time spent collecting water, sanitation practices, and satisfaction with services. Qualitative data were collected through key informant interview and FGD guides, while structured observation checklists were used to assess the functionality and condition of water infrastructure. Secondary data from programme reports and financial records were also reviewed.

### Data analysis

Quantitative household survey data were collected via KoBoCollect, uploaded to a secure server, and subjected to ongoing quality checks. After fieldwork, data were cleaned and analysed using IBM SPSS Version 27. Descriptive statistics summarized key indicators, while inferential analyses (including chi-square tests, t-tests, and odds ratios) assessed associations and measured change over time against WaterStarters baseline values. Qualitative data from key informant interviews and focus group discussions were transcribed, translated where necessary, and analysed thematically using NVivo. A structured coding framework guided the identification of recurrent themes and explanatory insights aligned with the evaluation objectives. Financial analysis assessed the viability of the franchise model through cost classification, break-even calculations, ROI, and NPV estimations, alongside cash flow and revenue trend assessments. Data triangulation across methods, respondent types, and data sources enhanced validity and provided a comprehensive understanding of programme outcomes, implementation fidelity, and contextual drivers of observed changes.

### Ethical considerations

Ethical approval was obtained from the Amref Health Africa Ethics and Scientific Review Committee (ESRC), with additional clearance from county authorities. Informed consent was obtained from all participants, and participation was voluntary. Data were anonymized and confidentiality strictly maintained. The evaluation team was trained in research ethics, cultural sensitivity, and safeguarding of respondent welfare.

## Results

### Respondent and household characteristics

The majority (71%) of the respondents were women which reflects the gendered roles in water collection and household management. Ages ranged from 16 to 80, with an average age of 38 years. Most respondents (75.3%) were married, and nearly half identified as spouses of household heads, while a third reported being heads of household themselves. Educational attainment varied, with 40.6% having completed secondary education, 29.6% primary, and 20.6% tertiary. Notably, 9.1% had no formal education, with gender disparities evident - 10% of women lacked formal education compared to 7% of men. These trends mirror national surveys highlighting persistent education gaps, particularly among women in arid and semi-arid regions like Kajiado. Household sizes averaged five members while 62% of households had at least one child aged under five years. The socio-economic profile of surveyed households revealed a predominance of informal, agrarian livelihoods. Livestock keeping was the main source of income for 43% of households, consistent with Kajiado’s pastoralist socio-economic profile. Small-scale trade and business activities accounted for 19.8%, especially in peri-urban areas such as Kibiko. Other income sources included casual labour (15.2%), formal employment (14.7%), and crop farming (5.6%).

The International Wealth Index (IWI) was used to measure household wealth; this is a standardized measure based on assets that was created by the Global Data Lab. The index is based on 12 indicators such as consumer durables, accessibility of electricity and better water, and housing conditions such as flooring type, sanitation, and the numbers of the sleeping rooms. Global principal component coefficients were used to weight each variable in order to get a composite IWI score (0100), which was then utilized to define households as five wealth quintiles. Figure 1 indicates that there were large disparities in wealth distribution between the five sites of the project. Kibiko turned out to be the richest location, boasting the 46 percent of households in the highest quintile and only the 8 percent in the lowest one. Unlike the other two, Orinie was the poorest, and 45% of the households in the area were in the lowest quintile with none being in the highest quintile. Kumpa and Naserian also had concentration in the lower two quintiles which is an indicator of economic vulnerability. Kima was more evenly distributed with 30 percent of households falling in the middle quintile and this indicated middle overall levels of wealth. These differences confirm the significance of location particular targeting approaches and the necessity of adjusting water access and pricing frameworks to the economic realities of every community.

**Fig 1:**
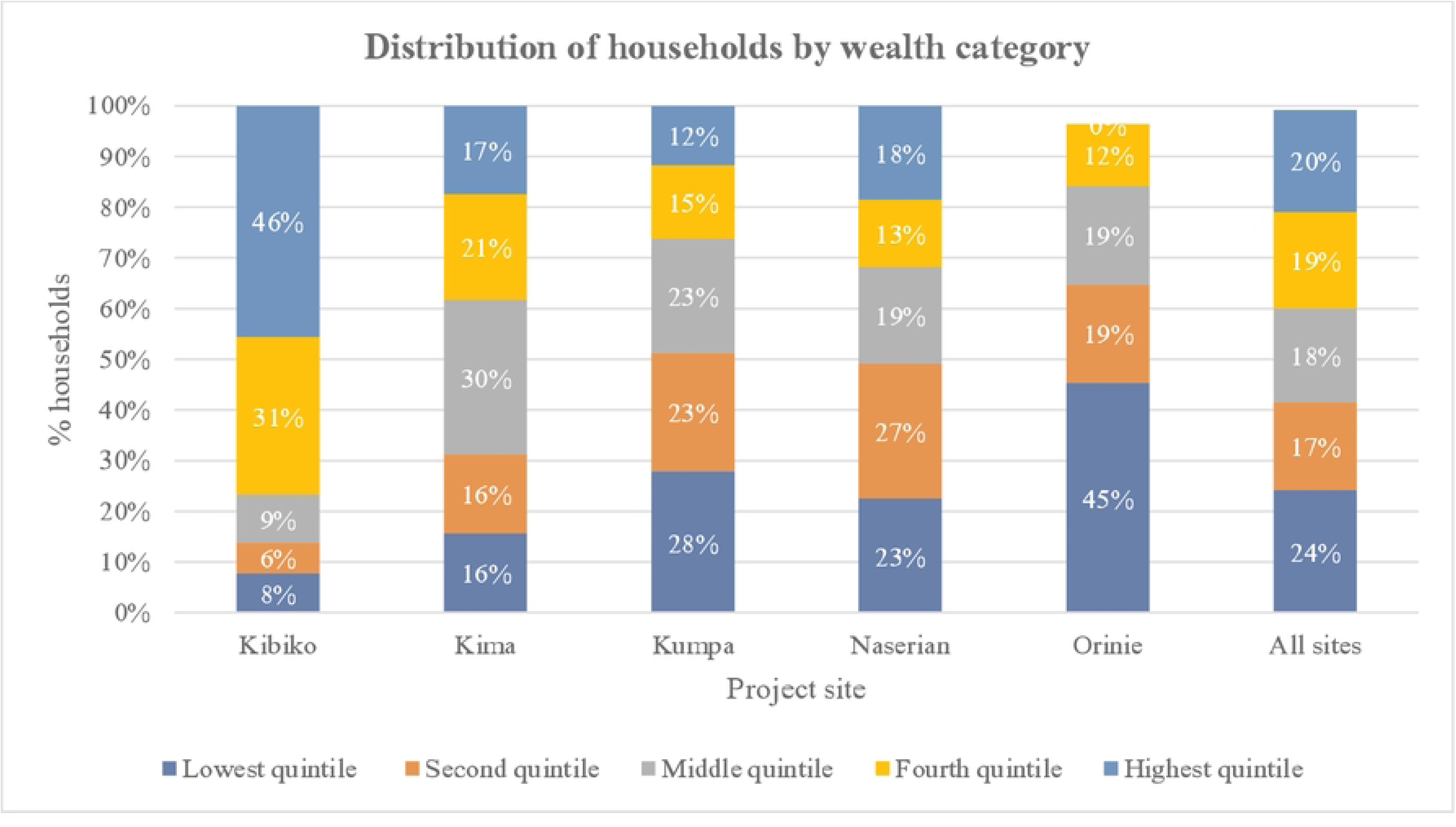
Distribution of households by wealth category.

### Stakeholder reception of the franchise model

The study found that the WaterStarters franchise model was broadly well received across stakeholder groups, including county officials, franchisees, and community members. Stakeholders commended the model for its participatory design, co-financing approach, and strong alignment with county development priorities - particularly around climate-smart infrastructure and public-private partnerships. Government officials highlighted the model’s potential to strengthen accountability and promote a sense of ownership among franchisees. The use of solar-powered pumps, prepaid meters, and real-time digital dashboards was widely appreciated for enhancing transparency, improving cost efficiency, and boosting operational resilience.

Despite this broad support, the study identified gaps in community awareness of the franchise structure, with some residents assuming the water systems were government-owned. This misconception may affect long-term payment compliance and expectations around service provision. Additionally, concerns were raised about the governance and financial management capacity of franchisees, underscoring the need for ongoing training and institutional support.

Overall, satisfaction with system management was high, with 95% of households reporting they were satisfied or very satisfied. A chi-square test confirmed a statistically significant association between project site and satisfaction levels (χ²(20) = 119.285, p < .001), highlighting the influence of site-specific factors on user experience and programme success.

### Access to safe and affordable water

The WaterStarters Programme has gone a long way in enhancing availability of safe, reliable and affordable water in project locations, and franchisee run water points are now the most common source of drinking water among a surveyed 59 percent of households across the project sites. Coverage was also maximal in Naserian (90%) and Orinie (79%), and uptake in Kibiko 20%) was trailing behind because of the availability of other formal utilities and potential underreporting associated with overlapping infrastructure. The perceived increase in water availability over the last 35 years were common, with the WaterStarters Programme being the most commonly attributed in Naserian, Orinie and Kumpa. The respondents had to walk miles to obtain water as one respondent remarked; *“We used to walk for hours to fetch water. Now it’s just nearby—and it’s clean.”*

The expanded service reach was particularly evident among lower-income households, implying a strong pro-poor orientation. 74% of households in the lowest wealth quintile relied on WaterStarters services, compared to just 31% in the highest quintile. A chi-square test confirmed a significant association between wealth and water source (χ²(32) = 425.12, p < .001), suggesting the model is effectively reaching underserved populations.

Progress along the JMP drinking water ladder reinforced these findings. Several sites saw notable shifts from limited or unimproved sources to safely managed or basic services. In Kibiko, safely managed access increased from 24% to 83%, while similar gains were recorded in Naserian and Kumpa. Affordability and improved convenience were key drivers of this progress, particularly in areas with previously distant or unsafe water options.

These results underscore the value of franchise-managed delivery as a scalable, equity-driven solution for expanding water access in underserved and resource-constrained settings. Still, barriers remained. Distance was the most cited reason for non-use, especially in Orinie, Kibiko, and Kima. Affordability was a concern in Naserian and Kumpa, while other households opted for pre-existing sources deemed satisfactory. Notably, 94% of WaterStarters users expressed satisfaction with water quality, compared to 84% among non-users (a statistically significant difference: p = 0.029).

### Water pricing and affordability

An overwhelming 88% of households reported paying for water services, with the highest payment rates observed in Kumpa (95%) and the lowest in Naserian (76%). Among those who paid for water, more than half (52%) cited a WaterStarters franchise as their source - though this may be an undercount in peri-urban Kibiko, where many users were unaware that their piped connections are part of the WaterStarters network. WaterStarters kiosks consistently offered the most affordable and predictable pricing - fixed at KSh 5 per 20-litre jerrican. In contrast, households purchasing from informal vendors paid significantly more, with costs ranging up to KSh 60 per jerrican. Piped water connections, while more convenient, attracted variable tariffs and surcharges. Despite this, nearly 80% of respondents rated kiosk water as affordable, reinforcing the model’s appeal as a pro-poor solution.

Affordability challenges were reported by 26% of all households, but this dropped to 18% among WaterStarters users, confirming the model’s relative accessibility. Still, site and wealth-level disparities persisted. In Orinie (with the highest proportion of low-income households), 40% reported struggles with water costs. A socio-economic gradient was evident, with 29% of households in the lowest wealth quintile reporting occasional inability to pay, compared to just 9% in the highest quintile.

A majority of households (55.3%) were unwilling to pay higher tariffs for WaterStarters services, highlighting widespread price sensitivity. Only 39.1% expressed willingness to pay more, with Kibiko showing the highest support (64.7%) and Kima the lowest (23.5%). Qualitative insights linked willingness to service reliability and lack of alternatives, while reluctance stemmed from financial hardship. In many cases, continued usage was driven by necessity rather than choice, raising equity concerns around future tariff adjustments without protective measures for vulnerable groups.

When unable to pay, households employed a mix of coping strategies. Over half reduced their water use, particularly in Naserian and Kibiko. Others turned to neighbours, free boreholes, or unprotected sources - raising public health concerns. A quarter admitted skipping hygiene practices, especially in Orinie and Kumpa. These coping mechanisms, while adaptive, expose vulnerabilities and highlight the need for targeted social protection to ensure water security for the poorest.

### Operational reliability

The WaterStarters franchise model has significantly enhanced the reliability of water systems across project sites, with stakeholders consistently reporting fewer breakdowns and faster response times. Franchisees and local officials noted that service interruptions are now rare and typically resolved within 72 hours. Routine preventive maintenance, regular infrastructure inspections, and real-time monitoring have been central to this operational consistency. Franchisees described proactively checking for wear and tear, while automated dashboards and early warning systems flagged anomalies in usage patterns, enabling rapid responses. Communication was further streamlined through WhatsApp groups linking franchisees, county officials, and technicians, ensuring swift coordination when issues arose.

Survey data echoed these stakeholder accounts. Among the 575 households relying primarily on WaterStarters-supported systems, 88% reported no mechanical breakdowns in the past year. For the 34 households that did experience issues, the average repair time was just 2.74 days. Residents frequently highlighted the speed and reliability of repairs, with one community member stating, *“Now, if something happens, it’s fixed in a day. Before, we could go many days without water.”*

Confidence in the model’s sustainability remains high, with early 98% of users believing the system would continue functioning after Amref’s exit. However, a minority expressed concern about future privatization, cost hikes, or management lapses—underscoring the importance of robust governance and community engagement to safeguard long-term trust and equity in water access.

### Effects on livelihoods and climate resilience

The WaterStarters model has catalysed a transformation in how water is used within households, shifting it from a scarce resource reserved for drinking and cooking to an enabler of hygiene, productivity, and economic opportunity. With improved access, many families now use water more freely for cleaning, bathing, and household gardening. The availability of consistent water supply has also spurred a wave of income-generating activities (IGAs). Across project sites, 62% of households that reported improved water access said they had initiated or expanded an IGA - most notably in farming, poultry rearing, and livestock keeping. The rise of microenterprises such as water vending, car washes, and laundry services in peri-urban centers like Kumpa and Kibiko further illustrates how water infrastructure is enabling local entrepreneurship.

Significantly, lower- and middle-income households were the most responsive to these opportunities. A statistically significant relationship (χ²(4) = 11.205, p = .024) shows that poorer households were more likely to start IGAs following water improvements - suggesting that water access can help close livelihood gaps. Women, in particular, have been at the forefront of this shift. Many are creatively repurposing household wastewater to sustain backyard vegetable plots, bolstering food security and income through the sale of surplus produce. In the traditionally pastoralist communities, reliable water has sparked new interest in small-scale farming. Community leaders and organizations are experimenting with climate-smart agriculture reflecting the model’s potential to strengthen climate resilience.

*“Recently we’ve seen very good progress… many people are now trying farming. Near here, we have two farms run by women groups. Even we (local CBO), have started exploring farming for sustainability. We’ve fenced off a three-acre plot for horticulture and hope to see good outcomes.”*

### Effect on children’s education

Access to safe and reliable water through the WaterStarters Programme has emerged as a critical enabler of educational continuity, hygiene, and dignity for school-aged children— particularly girls. By combining water infrastructure improvements with school-based health and hygiene interventions, the programme has helped tackle persistent barriers to school attendance, including the burden of domestic water collection, frequent illness from unsafe water, and inadequate menstrual hygiene facilities.

Quantitative data showed that 14% of households reported increased school attendance as a direct result of time saved from water-fetching responsibilities. This impact was especially evident in areas like Kumpa and Naserian. A head teacher in Kumpa reported that school enrolment had increased from 600 to over 950 pupils since the introduction of the new water system, attributing the surge to improved water access. Qualitative accounts echoed these trends. Community leaders and educators consistently linked the availability of nearby water to better attendance, classroom participation, and school morale. Girls, in particular, have benefitted from reduced domestic burdens, allowing them to attend school more consistently. *“Before, children were told to skip school to fetch water or tend to livestock. Now, with water close by, they can focus on learning,”* noted one community leader.

For adolescent girls, the changes have been especially transformative. With the introduction of private washing areas, sanitary disposal facilities, and reliable handwashing stations, menstrual hygiene management has improved significantly. These improvements have enhanced not only attendance but also confidence and dignity, enabling girls to participate in school without interruption or stigma.

### Time savings from improved water access

Improved water infrastructure under the WaterStarters Programme has led to significant time savings for households across project sites - time that is now being reclaimed for education, income generation, and caregiving. The JMP defines 30 minutes or less for a round-trip to fetch water as the threshold for basic access. Across four of the five WaterStarters sites, the proportion of households meeting this threshold rose markedly. Kumpa led the gains, improving from 44% at baseline to 88% at midterm, followed by Kibiko (67% to 89%) and Kima (55% to 74%). Even Orinie, where access remains limited, nearly doubled its coverage (14% to 27%).

Nearly half of households reported saving between 30–60 minutes per round trip, while in Kima, 21% reported saving up to two hours. The freed-up time was put to good use: rest and leisure (73%), housework (65%), income-generating activities (48%), caregiving (47%), and community involvement (46%) were the most commonly reported uses. With less time spent collecting water, many women reported greater participation in social networks and personal wellbeing. One woman in Kibiko explained, *“Now with water nearby, I can attend and participate fully in our chama* [informal investment club] *meetings.”* The findings highlight not only household-level improvements but also broader gains in women’s empowerment and social inclusion.

### Change in sanitation coverage and practices

The WaterStarters programme has driven notable improvements in household sanitation coverage and hygiene practices across its five implementation sites. Between baseline and midterm evaluations, latrine access increased substantially: Kumpa rose from 55.9% to 84.2%, Orinie from 59.7% to 71.3%, Naserian from 79.0% to 92.5%, and Kima from 88.9% to 95.7%. Kibiko maintained near-universal coverage at 99%. These gains were widely attributed to the hygiene promotion efforts of Community Health Promoters (CHPs) supported by county public health departments.

Nonetheless, 11.2% of households still lacked their own toilet or latrine. Of these, 76% practiced open defecation and 22% shared facilities - signalling persistent gaps that demand sustained investment and behaviour change initiatives.

Further analysis using the WHO/UNICEF JMP sanitation ladder revealed site-specific improvements in sanitation quality. Kibiko saw safely managed sanitation rise from 7% to 32%, while open defecation fell from 8% to 1%. Kima recorded a dramatic rise in basic sanitation (15% to 65%) and a corresponding drop in limited sanitation (59% to 19%). Kumpa achieved a reduction in open defecation from 48% to 16%, alongside modest gains in basic services (1% to 35%). Naserian posted the most dramatic increase in basic sanitation (from 29% to 79%) and reduced open defecation to 8%. Orinie made progress but remains the most challenged site, with 29% still practicing open defecation and safely managed sanitation negligible.

The safe disposal of children’s faeces remains a critical concern. While 58% of households reported safe practices (e.g. rinsing or placing faeces into toilets or latrines) wide disparities exist. Naserian led with 85% compliance, followed by Kibiko (69%). Kima lagged at 37%, with 31% of households still burying faeces - an unsafe practice by JMP standards. Other unsafe methods included disposal in garbage (14%) and drainage systems (1%).

These findings reflect both the gains and ongoing challenges in sanitation service delivery. While the WaterStarters model has catalysed important improvements, open defecation and unsafe child faeces disposal remain critical issues requiring intensified behaviour change interventions.

### Effect on handwashing practices

The WaterStarters Programme’s impact on household hygiene reveals persistent disparities across sites, with most households lacking basic handwashing facilities. In Orinie, Kumpa, and Naserian, over 65% of households had no handwashing setup, while Kima led with 43% reporting basic hygiene services—attributed to active Community Health Promoters (CHPs). Kibiko followed at 29%, reflecting higher household water connectivity. However, many households still fell into the “limited hygiene service” category, lacking either soap or water, which undermines infection prevention.

While handwashing after defecation (90%) and before eating (80%) was widely practiced, less than a quarter of households reported handwashing before feeding a child (21%) or after changing a baby (15%) - critical gaps for preventing disease transmission. CHPs played a central role in driving behaviour change, with over 85% of households citing home visits as the main source of hygiene education. Community meetings also contributed, especially in rural areas like Orinie and Naserian.

### Health gains

The WaterStarters Programme yielded promising public health benefits, particularly in curbing diarrhoeal disease. Survey data across five sites showed just 1% of households reported diarrhoea in the prior two weeks, down sharply from baseline rates that reached as high as 23.5% in some areas. Household perceptions echoed the trend. In Naserian and Orinie, virtually all respondents observed a decline in diarrhoea, while Kumpa and Kima reported more modest improvements. Kibiko showed the least perceived change, with many respondents unsure, suggesting either lower programme visibility or weaker outcomes.

Health facility data added further insight. In Kumpa, reported diarrhoea and eye infections declined between 2023 and 2024, especially among young children. Kima, however, saw a rise in facility-reported diarrhoea cases, likely reflecting better care-seeking behavior promoted by active CHPs, rather than worsening disease. These findings highlight the Programme’s impact while also signalling the importance of continued monitoring and targeted interventions, particularly for the most vulnerable populations.

### Financial performance of waterstarters franchisees

Financial performance analysis across the five pilot sites reveals both promise and pressure points, underscoring where the model works, where it falters, and how it could be optimized.

Of the five sites reviewed only one emerged with a positive financial outcome under the original capital structure. Kibiko posted a Net Present Value (NPV) of KES 1.1 million and a Return on Investment (ROI) of 14.3%, signalling that it not only covers costs but can generate surplus value. Its success was attributed to a balanced start-up cost of KES 7.7 million, steady revenue inflows, and a robust technical backbone. The other sites, however, told a different story. Kima, despite being the most heavily capitalized at KES 11.6 million, recorded an ROI of –80.8%, with losses driven by slow revenue growth and high debt servicing costs. Orinie, with the smallest capital outlay, fared no better (an ROI of –76.1%) suggests that limited investment did not shield it from poor financial returns. Kumpa and Naserian posted moderate losses but still remained below breakeven thresholds.

**Table 1:** Capital investment analysis summary.

| Site | Total Investment (KES) | NPV (KES) | ROI (%) |
| --- | --- | --- | --- |
| Kibiko | 7,702,218 | 1,103,917.64 | 14.33 |
| Kima | 11,614,508.97 | -9,381,438.39 | -80.77 |
| Kumpa | 8,860,352.14 | -3,643,713.75 | -41.12 |
| Naserian | 9,240,288.09 | -2,551,788.8 | -27.62 |
| Orinie | 3,602,560.44 | -2,741,330.11 | -76.09 |

These findings point to a common constraint: a heavy loan component (42.5%) in the initial capital mix proved unsustainable across most sites. Revenue generation in rural settings was simply too slow and too uncertain to support large debt obligations early in the franchise lifecycle. As a result, liquidity constraints and delayed breakeven timelines hampered financial stability, particularly at sites with high Capital Expenditure (CapEx) and seasonal fluctuations in demand.

To test alternative pathways to viability, a Minimum Viable Product (MVP) scenario was modelled for each site, featuring a reduced capital input and rebalanced financing mix. The results were striking: all five sites returned positive NPVs, and ROI values, while modest, turned from negative to positive. Kima, previously the worst performer, showed the most dramatic turnaround. Under the MVP scenario, with a lower loan share (29%) and higher grant allocation (51%), it posted the highest ROI of 4.98% and an NPV of KES 431,108. This not only validated the potential of the site under leaner conditions but also highlighted the outsized impact of capital structure on early-stage viability. Kibiko again demonstrated solid performance, returning an ROI of 1.14% on a trimmed investment of KES 6.6 million. The other sites (Kumpa, Naserian, and Orinie) posted ROI figures ranging from 0.48% to 2.57%, each showing signs of financial health when debt exposure was reduced and grant support bolstered. These improvements confirm that right-sizing capital inputs (leaner, smarter capital) and recalibrating the loan-to-grant ratio can transform financial trajectories even in challenging rural markets.

**Table 2:** Minimum viable product (MVP) scenario.

| Site | MVP Investment<br>(KES) | Loan<br>(%) | Grant<br>(%) | Equity<br>(%) | NPV<br>(KES) | ROI<br>(%) |
| --- | --- | --- | --- | --- | --- | --- |
| Kibiko | 6,623,907 | 35 | 44 | 21 | 75,792.24 | 1.14 |
| Kima | 8,652,809 | 29 | 51 | 20 | 431,108.50 | 4.98 |
| Kumpa | 6,805,990 | 44.9 | 40.99 | 14.11 | 174,962.90 | 2.57 |
| Naserian | 6,098,590 | 23 | 55 | 23 | 29,984.20 | 0.49 |
| Orinie | 2,990,125 | 31 | 51 | 18.00 | 14,345.09 | 0.48 |

In view of the foregoing, what then represents the optimal balance for financing rural water enterprises in the context of Kajiado?

Synthesizing scenario results and repayment data, the analysis proposed an optimal capital structure comprising 14.2% loan, 66.2% grant, and 19.7% franchisee equity. This represents a sharp pivot from the original 42.5% loan share and directly responds to repayment challenges observed across the pilot cohort. The reduced debt load offers crucial breathing room during the high-risk start-up period, while the generous grant provision ensures projects can focus on operational stabilization before profitability. Yet the retention of a nearly 20% equity contribution from franchisees ensures local commitment and accountability - a key to long-term sustainability.

Overall, the findings point to a key conclusion that the WaterStarters model, in its initial form, pushed the limits of financial realism in low-income settings. While technically sound and socially impactful, the capital-heavy approach placed undue strain on early-stage operations, jeopardizing viability in most sites. However, this study reveals that with modest recalibrations (specifically, reducing loan exposure and increasing grant support) financial sustainability is within reach. The MVP and optimal financing scenarios provide a compelling blueprint for future rollouts: smart capital, tailored to local cash flows, can unlock not just water access but financial resilience.

Thus, based on cross-site scenario analysis, the optimal capital mix comprises 66% grant, 20% franchisee equity, and 14% loan. This configuration balances affordability, sustainability, and franchisee ownership while minimizing early liquidity stress.

### Scaling potential and determinants of success for the waterstarters franchise model

Designed to overcome the persistent shortcomings of traditional community-managed water schemes, the model embeds local ownership and entrepreneurial investment to improve accountability and sustainability. Technology plays a central role - prepaid meters and real-time dashboards have strengthened revenue collection and operational oversight, enhancing the model’s appeal to both investors and county governments. Demand for replication is growing. Community leaders and residents from surrounding areas have expressed interest, and several county officials are keen to adopt the model, although limited fiscal space remains a constraint to public co-financing. Household willingness to pay for piped water connections is on the rise across all sites. However, scaling will require tackling several challenges. These include refining the financing structure (e.g., concessional loans, flexible repayment), diversifying revenue streams (e.g., agriculture, institutional supply), and tailoring the model to serve lower-density or low-income settings. Success at the franchise level depends on a mix of factors. High-performing franchisees were locally embedded, entrepreneurial, and operated in denser or mixed-use areas where demand is high and consistent. Franchisee capability emerged as both a risk and an opportunity. While some demonstrated strong business acumen, others needed support in budgeting, preventive maintenance, and loan management. Ongoing technical assistance, peer learning, and structured capacity-building interventions will be critical for enabling franchises to operate as sustainable enterprises.

### Conclusion

The WaterStarters Programme has demonstrated strong potential as a sustainable and scalable solution for improving water access in underserved communities. By integrating a social franchise model, the programme combines private-sector efficiency with public health goals, overcoming challenges such as poor accountability and limited sustainability in traditional community-managed water systems. It has consistently delivered reliable services and high franchisee compliance, enhanced by solar-powered systems and digital tools like prepaid meters and real-time dashboards, which have improved revenue collection and service monitoring. Additionally, the programme has spurred wider development benefits, including increased food production, diversified income sources, and greater resilience to climate shocks, driven by irrigated agriculture and water-enabled micro-enterprises. However, challenges remain, particularly in financial performance, with early revenues falling below projections due to high capital costs and loan-heavy financing. Revised investment models with lower debt ratios and higher grants are suggested to improve sustainability. Some sites also face issues with low willingness to pay, inaccurate hydrological assessments, and gaps in franchisee financial management.

## Authors’ Contributions

Conceptualization: KO, KKi, MM, KKa

Methodology: KO, KKi, MM, KKa

Formal analysis: KKa, SK, AM

Investigation (data collection): KKa, SK, AM

Writing – original draft: KKa, SK, AM

Writing – review & editing: KO, KKi, MM, GW, KKa

Supervision: KO

## Data Availability

All relevant data are within the manuscript and its Supporting Information files. However, the raw data supporting this study are not publicly available due to ethical considerations. However, anonymized datasets can be provided upon reasonable request. Quantitative data may be accessed by contacting the corresponding author listed in the manuscript.

---

**Fig 2:**
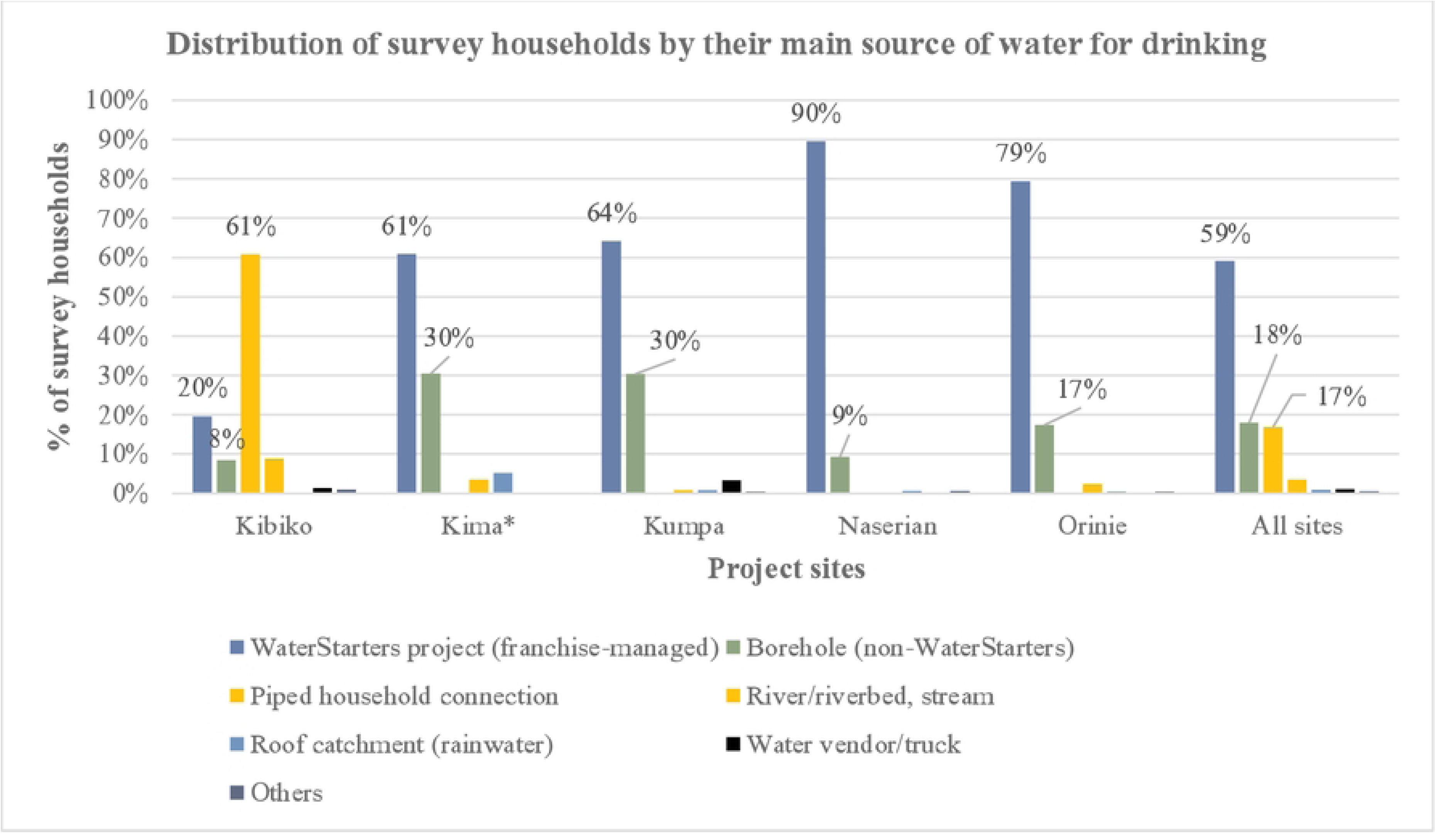
Distribution of survey households by their main source of water for drinking.

**Fig 3:**
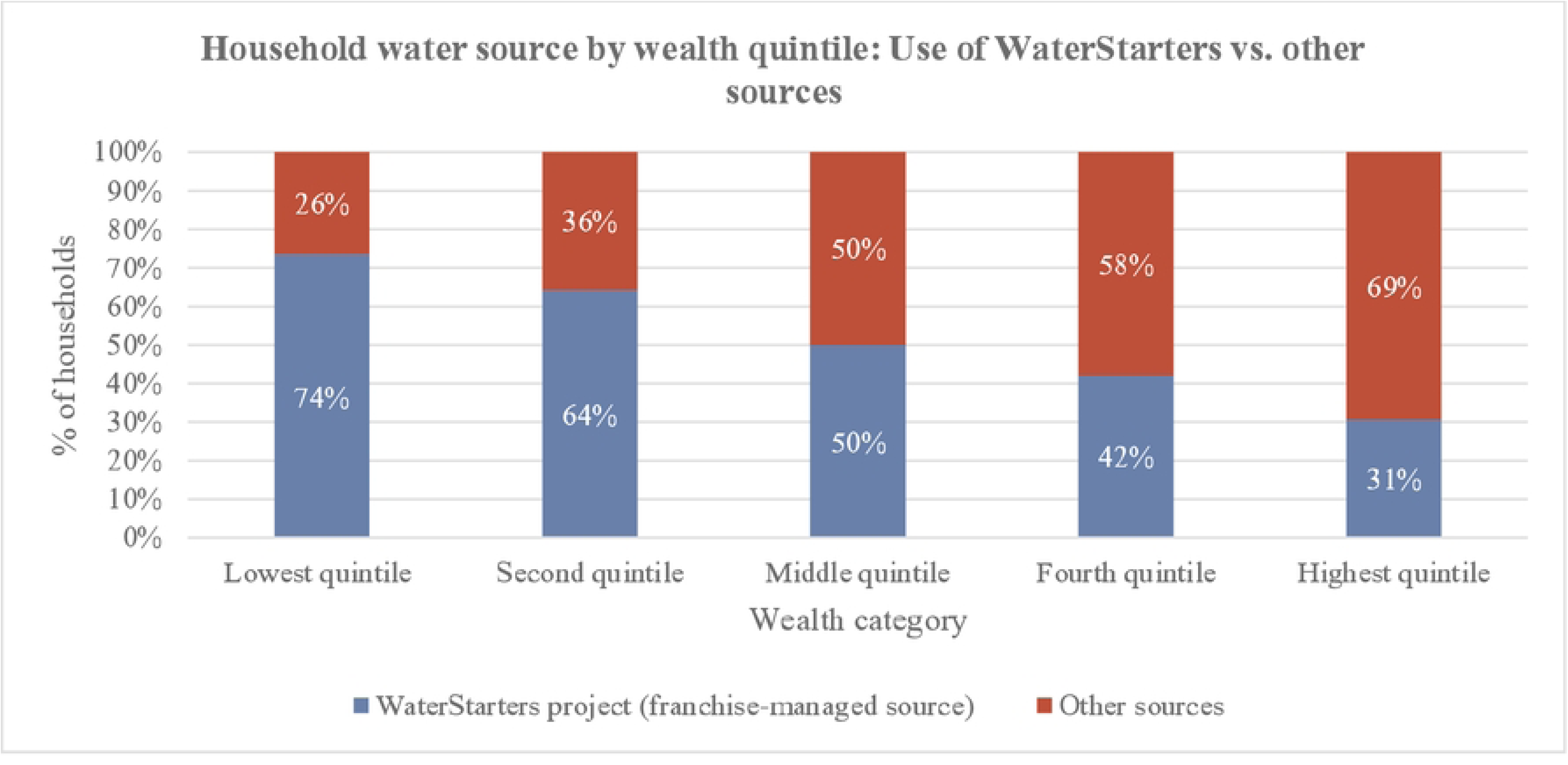
Household water source by wealth quintile: Use of WaterStarters vs. other sources.

**Fig 4:**
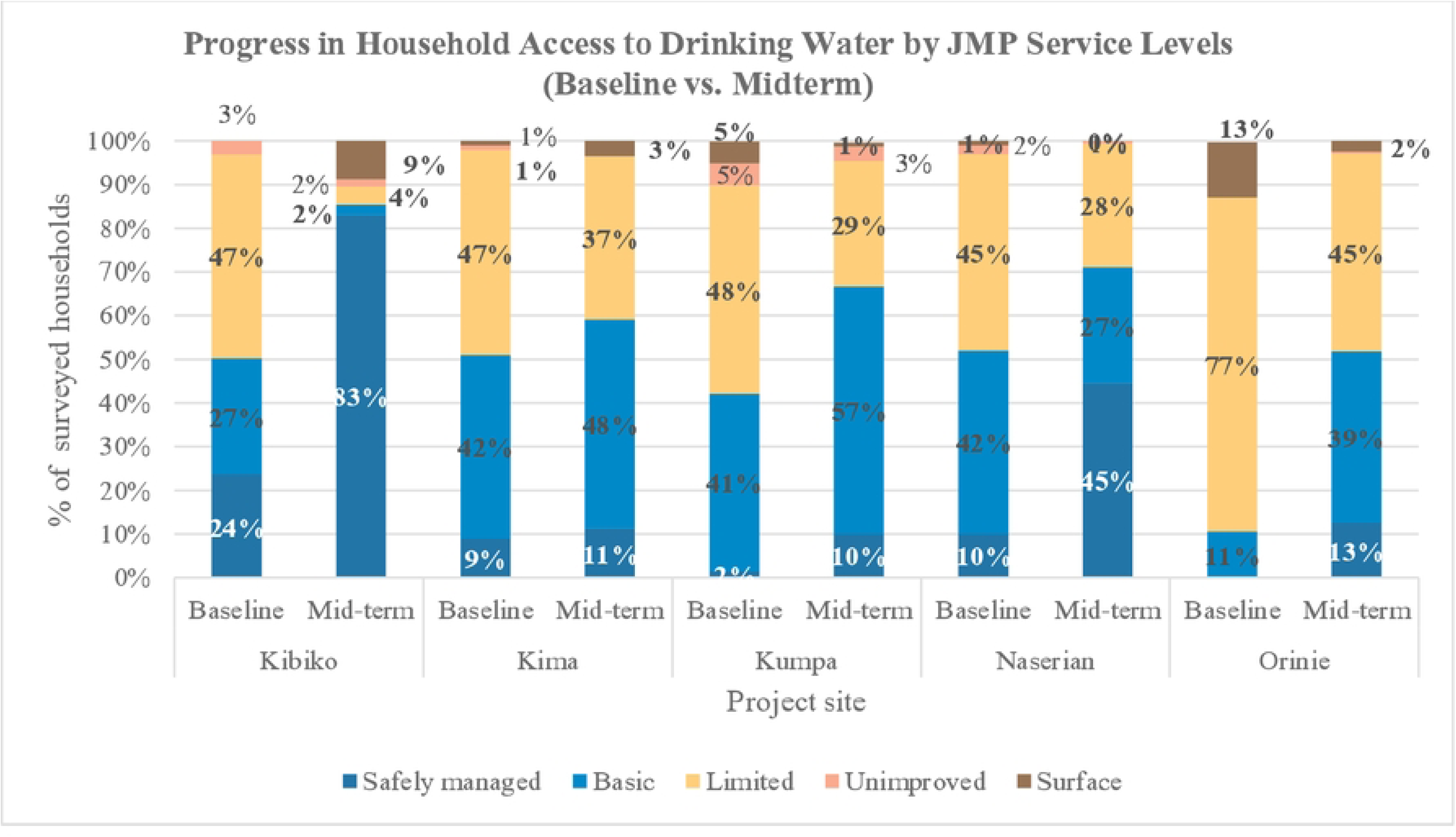
Progress in Household Access to Drinking Water by JMP Service Levels (Baseline vs. Midterm).

**Fig 5:**
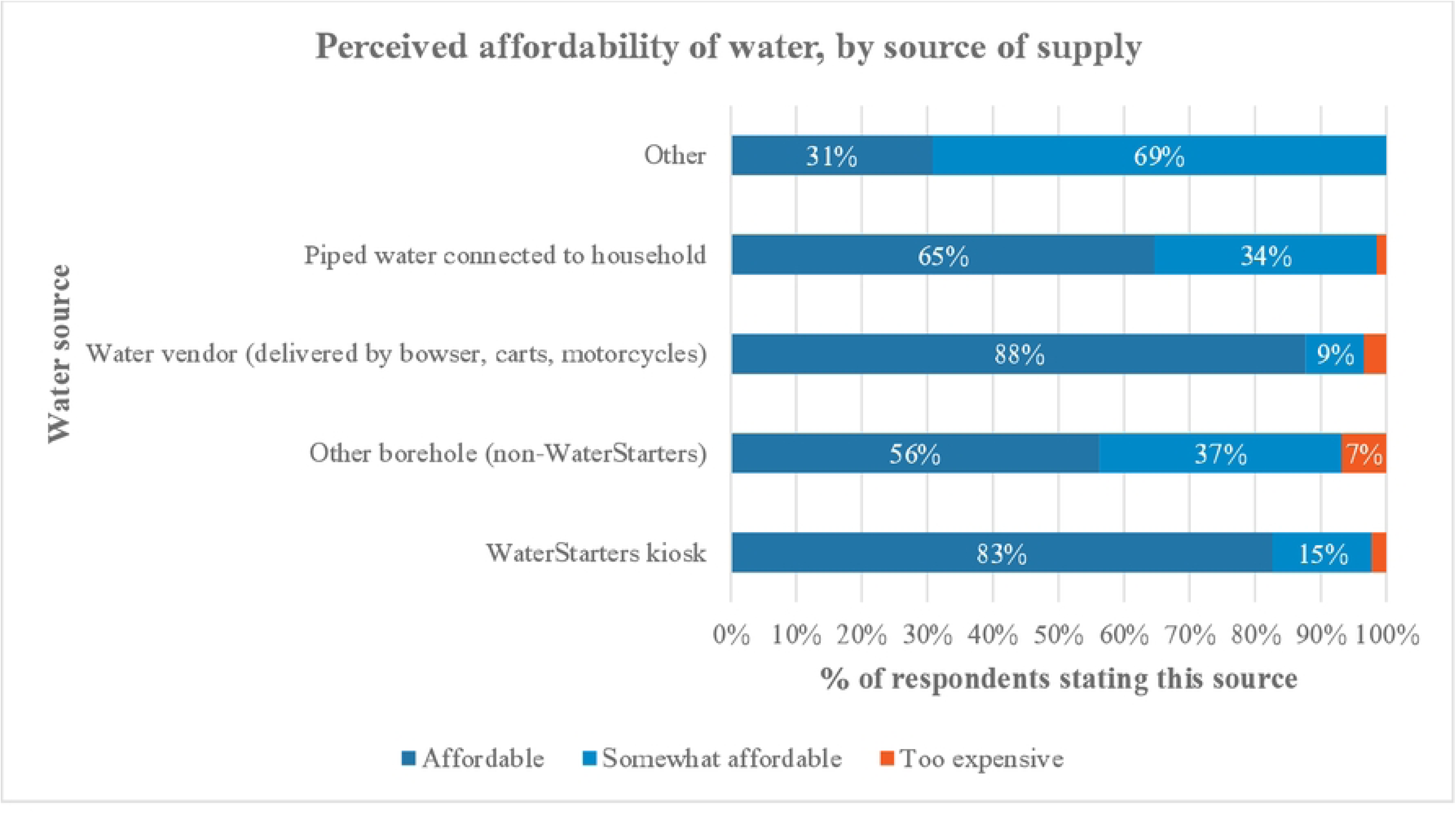
Perceived affordability of water, by source of supply.

**Fig 6:**
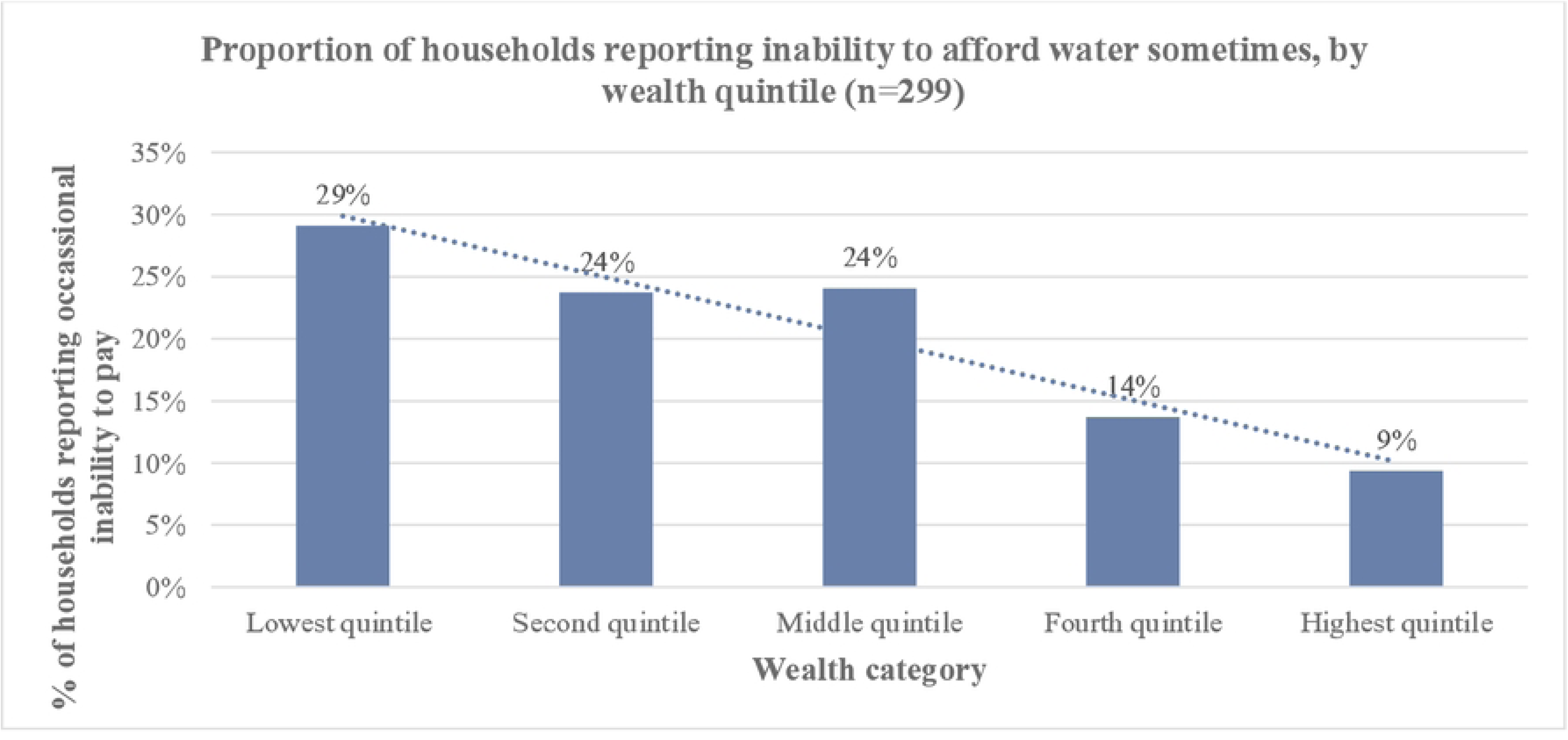
Proportion of households reporting inability to afford water sometimes, by wealth quintile.

**Fig 7:**
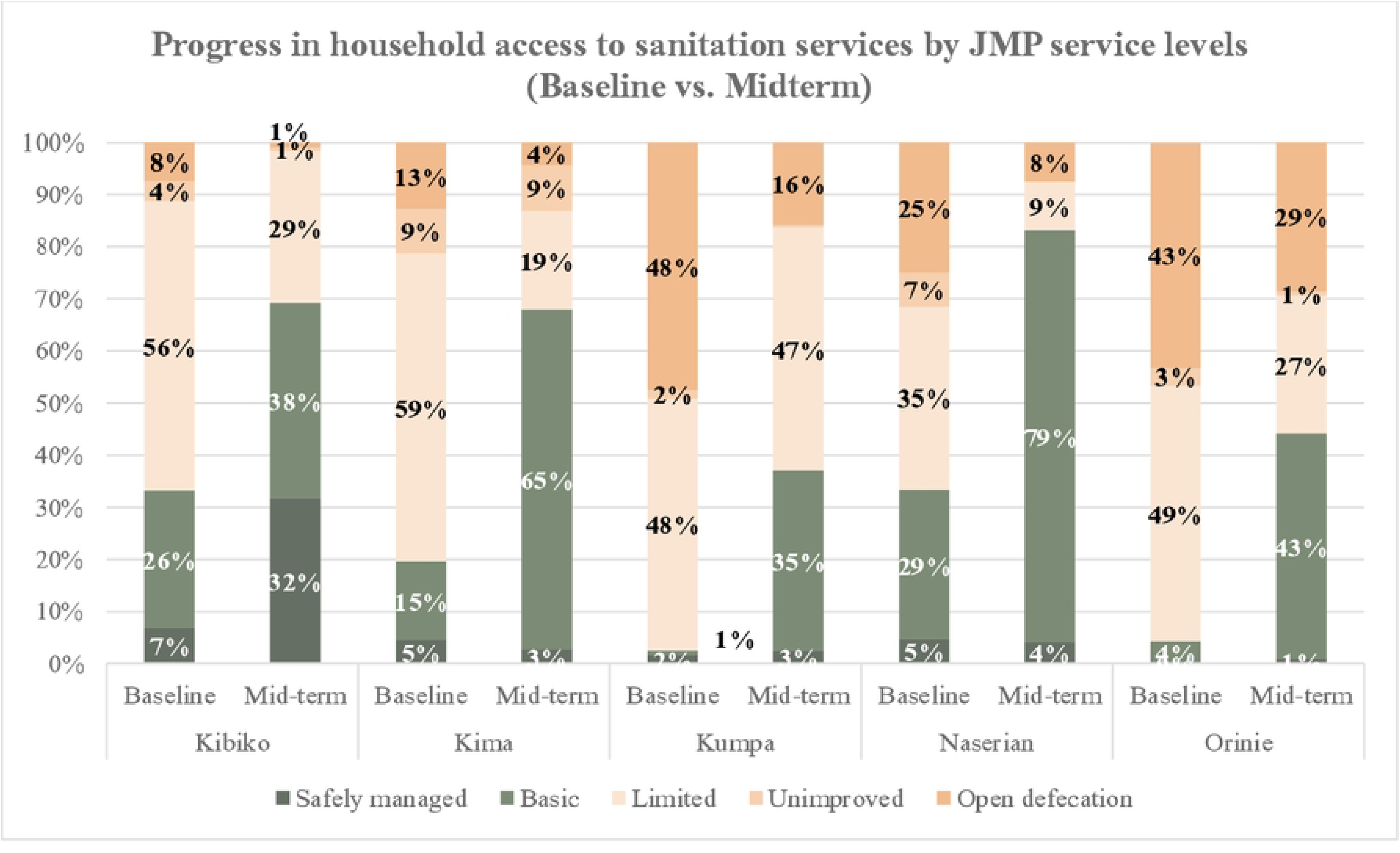
Progress in household access to sanitation services by JMP service levels (Baseline vs. Midterm).

**Fig 8:**
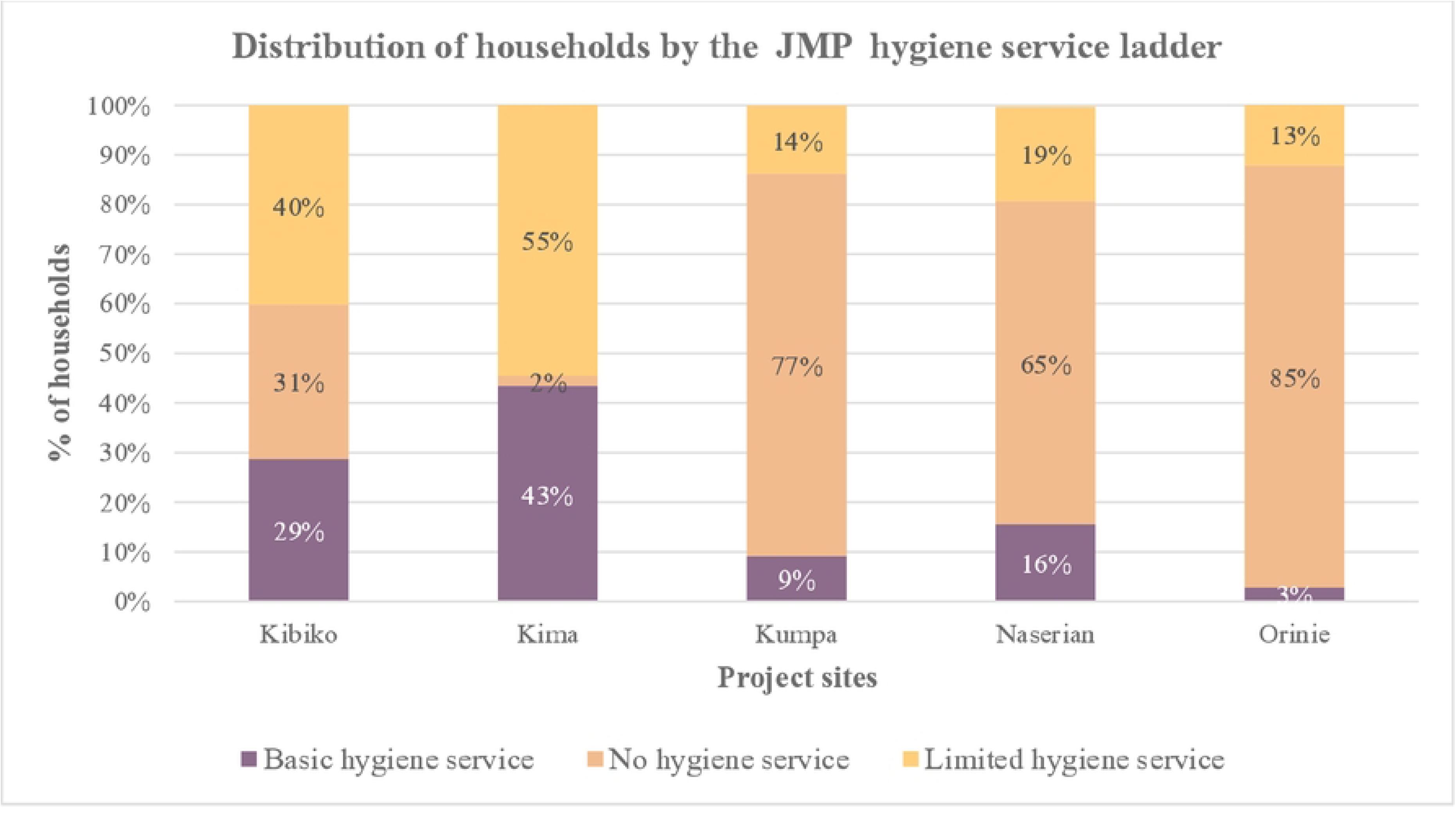
Distribution of households by the JMP hygiene service ladder.

**Fig 9:**
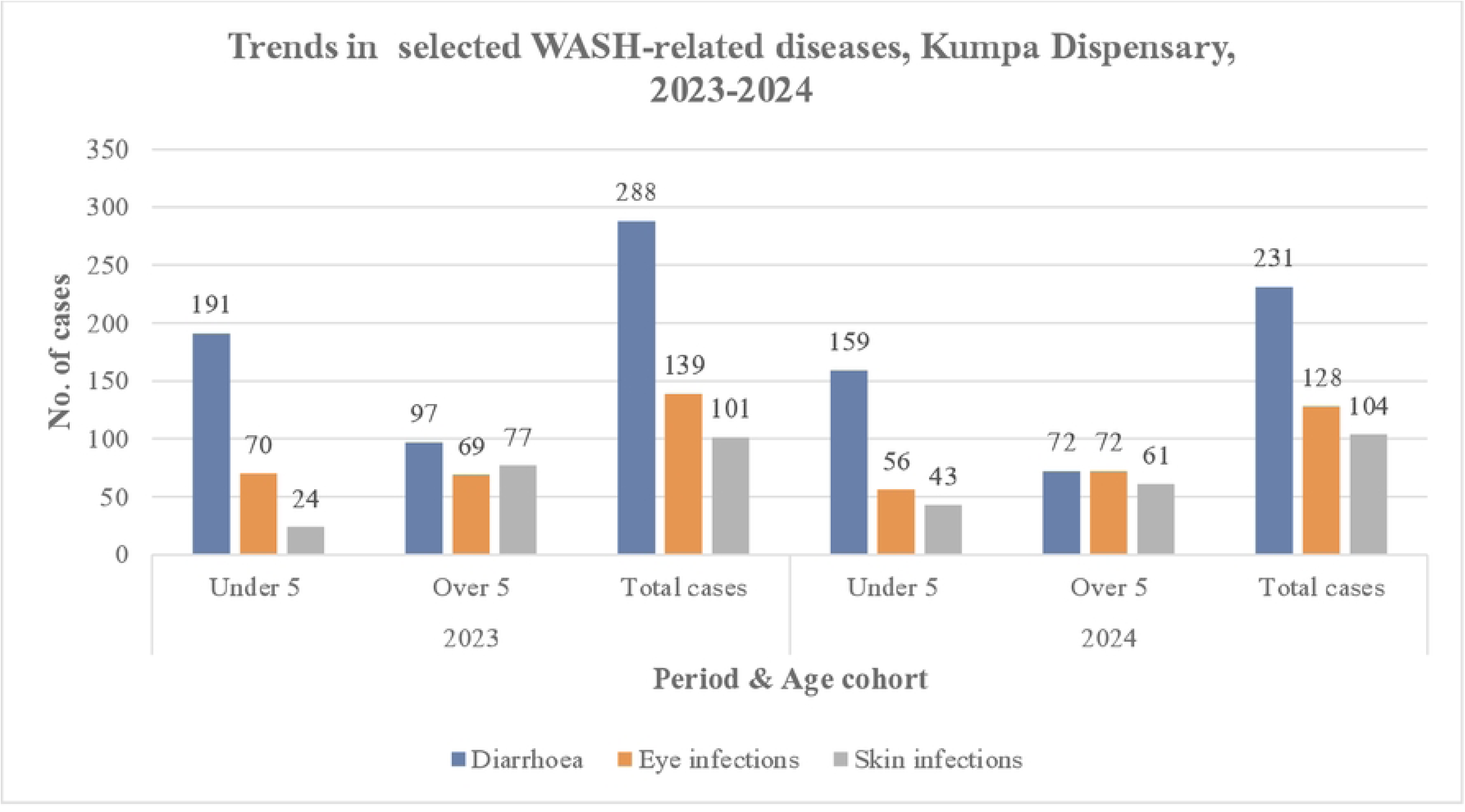
Trends in selected WASH-related diseases, Kumpa Dispensary.

**Fig 10:**
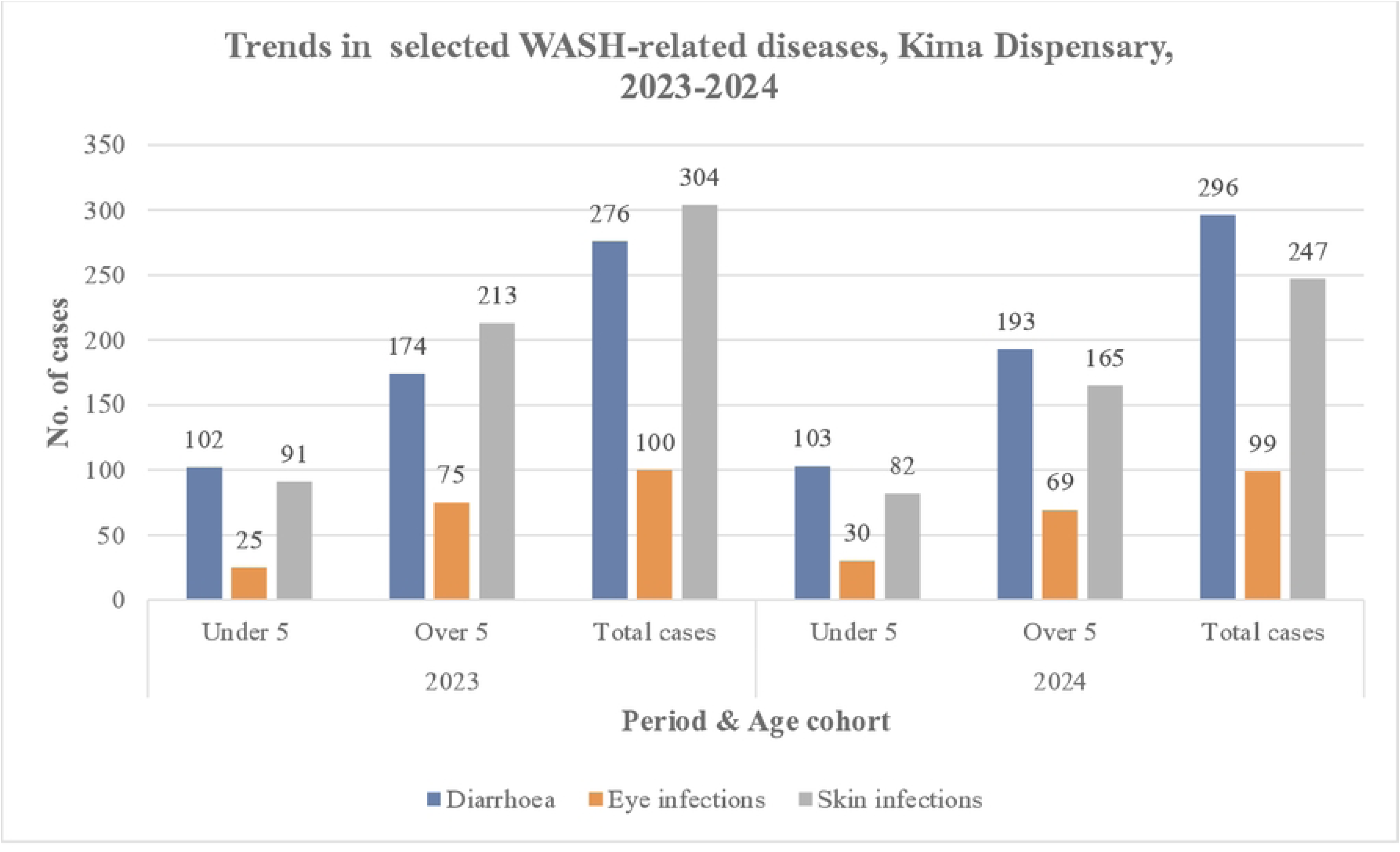
Trends in selected WASH-related diseases, Kima Dispensary.

